# A custom phenotypic profile for Fanconi anemia: Addressing gaps in existing disease annotations

**DOI:** 10.64898/2026.02.10.26346018

**Authors:** Evan Connelly, Bryan Laraway, Kathleen R. Mullen, Christopher J. Mungall, Melissa A. Haendel, Eric G. Hurwitz

## Abstract

Fanconi anemia (FA) is a rare genetic disorder of impaired DNA repair characterized by progressive bone marrow failure, congenital malformations, and cancer predisposition. Early identification of individuals with FA is critical for timely clinical management, yet phenotype-driven approaches to FA identification are hindered by inconsistencies in existing phenotypic profiles. We compared the Human Phenotype Ontology (HPO) annotations for FA in OMIM (215 terms across 22 complementation group entries) and Orphanet (106 terms in a single entry, ORPHA:84), quantifying overlap and anatomical system coverage. To address identified gaps, we developed a comprehensive custom HPO profile by extracting phenotypic terms from the entire Fanconi Cancer Foundation (FCF) Clinical Care Guidelines using OntoGPT, an LLM-based ontology extraction tool, followed by manual curation to ensure accuracy and clinical relevance. OMIM and Orphanet shared only 36 HPO terms (12.6% of their combined 285 unique terms), demonstrating substantial discordance. Our custom profile comprises 264 unique HPO terms, of which 161 (61.0%) are novel and not present in either existing source. The novel terms expand coverage particularly in musculoskeletal (39 terms, 23.8%), genitourinary (26 terms, 15.9%), limb (26 terms, 15.9%), head or neck (20 terms 12.2%), and digestive system (17 terms, 10.4%) phenotypes. Community-curated phenotypic profiles derived from clinical practice guidelines can substantially augment existing disease annotations. Our FA profile, the most comprehensive HPO-based phenotypic characterization of FA to date, is publicly available and provides a foundation for improved clinical decision support and EHR-based computable phenotyping that can accelerate diagnosis for individuals with FA. Furthermore, the LLM-assisted approach offers generalizable methods to improve the diagnostic odyssey for all rare diseases.

## Introduction

Fanconi anemia (FA) is a rare genetic disorder caused by pathogenic variants in genes encoding components of DNA damage repair pathways [1]. The disorder is characterized by progressive bone marrow failure typically presenting before age 10, congenital malformations affecting multiple organ systems, and a markedly elevated risk of hematologic and solid malignancies [2]. Because individuals with FA have hypersensitivity to DNA-damaging chemotherapy and radiation [3], timely diagnosis is essential to ensure appropriate hematologic monitoring, planning for bone marrow transplantation, and modified cancer treatment protocols.

The clinical presentation of FA is highly variable across the 22+ FA genes, even among individuals with variants in the same gene. Physical manifestations span virtually every organ system, including skeletal anomalies (particularly radial ray defects), short stature, skin pigmentation changes, genitourinary malformations, and characteristic craniofacial features. Due to phenotypic overlap with Vertebral, Anal, Cardiac, Tracheoesophageal fistula, Esophageal or duodenal atresia, Renal, upper Limb, and Hydrocephalus (VACTERL-H) association [4], the FA-specific PHENOS scoring system was developed as a clinical tool to identify patients requiring chromosomal breakage or genetic testing [5]. However, up to 40% of individuals with FA lack overt phenotypic anomalies, complicating clinical recognition [3]. The gold standard for diagnosis remains the chromosomal breakage test with diepoxybutane or mitomycin C [6], but this test is typically ordered only when FA is clinically suspected, creating a diagnostic bottleneck for individuals whose presentation does not fit the classic phenotypic pattern.

Identifying individuals with FA in electronic health record (EHR) data has been particularly challenging. Prior to October 2024, there was no specific International Classification of Diseases (ICD-10-CM) code for FA; the condition was subsumed under *other constitutional aplastic anemia* (D61.09). Although a Systematized Nomenclature of Medicine Clinical Terms (SNOMED CT) code for FA existed (30575002), its adoption in clinical practice has been limited. The introduction of ICD-10-CM code D61.03 for FA in October 2024 [7] represents an important advance, but the historical absence of specific coding means that retrospective identification of individuals with FA in EHR data requires phenotype-driven approaches.

The Human Phenotype Ontology (HPO) [8–10] provides a standardized, computable vocabulary for describing phenotypic abnormalities observed in human disease. The Monarch Initiative is an open-source consortium and platform that integrates gene, phenotype, and disease data across species to support translational research and clinical diagnostics [11,12]. HPO-based phenotypic profiles are maintained for thousands of rare diseases by two major resources: OMIM [13] and Orphanet [14]. These profiles serve as reference standards for computational phenotyping, clinical decision support, and variant prioritization in genomic diagnostics [10,15]. The Monarch Initiative integrates these and other resources, including gene, phenotype, and disease data across species, into a unified knowledge graph that enables semantic similarity-based analyses such as variant prioritization, patient profile matching, and phenotype-driven disease diagnosis [11,12]. For FA, 22 separate entries corresponding to individual complementation groups are maintained for content present in OMIM, while a single unified profile (ORPHA:84) reflects content in Orphanet. However, discordance between OMIM and Orphanet phenotypic profiles has been recognized as a challenge across rare diseases, and FA is no exception [14,16]. These resources differ in their curation approaches: OMIM annotates phenotypic features at the level of individual genetic subtypes based on published case reports, while Orphanet curates broader disease-level profiles based on expert review. Neither resource systematically incorporates the comprehensive clinical knowledge captured in disease-specific clinical practice guidelines [8].

The Fanconi Cancer Foundation (FCF) publishes and maintains Clinical Care Guidelines for FA [17], a regularly updated, evidence-based resource authored by FA clinicians for a clinical audience. These guidelines represent the most comprehensive synthesis of clinical knowledge about FA phenotypic manifestations, diagnosis, and management. Here, we present a community-curated HPO profile for FA derived from the complete FCF Clinical Care Guidelines, characterize its content relative to existing OMIM and Orphanet profiles, and demonstrate that clinical guidelines are a valuable and underutilized source for augmenting disease phenotype annotations. Building on the Monarch framework, our profile leverages HPO as the representational standard and is designed for integration into the Monarch knowledge graph, serving as a foundation for EHR-based computable phenotyping, semantic similarity-based diagnostics, and improved clinical decision support for FA.

## Methods

### Existing HPO profiles for FA

Existing HPO profiles for FA were obtained from the HPO annotation file (accessed January 2026) [8,9]. For OMIM, phenotype annotations were extracted for the current 22 FA complementation group entries (phenotypic series: PS227650) and the general FA entry (MIM:227650). Annotations were filtered to include only phenotypic abnormality terms and exclude negated annotations. The union of all OMIM FA entries was used as the combined OMIM profile. For Orphanet, the single FA entry (ORPHA:84) was used with the same filtering criteria. Term counts were deduplicated within each profile.

### Custom profile development from FCF clinical care guidelines

The custom FA phenotypic profile was developed by systematically extracting phenotypic information from the FCF Clinical Care Guidelines (5th Edition, 2020) [17]. The full text of each chapter was extracted from the https://fanconi.org website. Text was extracted directly; images were omitted, and tables and figures were adapted to flat text format suitable for processing by large language models.

OntoGPT [18], an LLM-based ontological extraction tool part of the Monarch Initiative ecosystem, was used to parse each chapter and identify candidate HPO terms. Each chapter was processed individually and duplicate entries were removed. For phenotypic terms identified by OntoGPT that could not be automatically mapped to HPO identifiers, CurateGPT [19], an LLM-based ontological curation tool, was employed to suggest appropriate HPO mappings. Notably, unmapped phenotypes have been flagged as candidates for HPO term requests, which in future iterations of the ontology could further refine FA phenotyping. The resulting candidate profile underwent manual review for clinical relevance. Each extracted term was evaluated in the context of its source chapter. Terms representing phenotypes directly attributable to FA were retained, while terms representing sequelae of treatment (e.g., chemotherapy side effects) or unrelated conditions discussed in the text were excluded. The final profile was deduplicated and filtered to include only terms that are descendants of *Phenotypic abnormality* (HP:0000118) in the HPO hierarchy.

### Profile Comparison

Profile overlap was assessed by comparing explicitly asserted HPO terms without transitive closure. Set operations were used to compute pairwise and three-way intersections among the OMIM, Orphanet, and custom profiles based on exact HPO term matches. This direct comparison quantifies the specific phenotypic assertions made by each database rather than semantic similarity through ancestral relationships in the HPO hierarchy. Terms present in the custom profile but absent from both OMIM and Orphanet were classified as novel.

### Anatomical system classification

Each HPO term in all three profiles was classified by anatomical system using the Histopheno API from the Monarch Initiative [20], which returns disease-phenotype annotation counts across 20 predefined top-level anatomical system categories for a given HPO term. For terms with multiple parent categories in the HPO hierarchy, system assignment was determined by clinical intuition based on the underlying pathology (e.g., leukemia was classified under neoplasm rather than blood abnormalities). This analysis was performed for each profile individually and for the subset of 161 novel terms unique to the custom profile.

### Use of large language models (LLMs)

Large language models, including OpenAI’s ChatGPT (GPT-4o, GPT-3.5) and Anthropic’s Claude (Opus 4.5, Opus 4.6, Sonnet 4.5), were used for editorial assistance with grammar, language, and word choice. All LLM suggestions were reviewed and approved by the authors; no LLMs were used to generate ideas.

## Results

### Discordance between existing FA phenotypic profiles

The OMIM combined FA profile contained 215 unique phenotypic abnormality terms across 22 complementation group entries. Individual OMIM entries varied substantially in size, from 5 terms (FANCJ) to over 50 terms (FANCD2), reflecting the phenotypic diversity across different pathogenic variant genes. The Orphanet FA profile (ORPHA:84) contained 106 unique phenotypic abnormality terms in a single unified entry. Despite both profiles describing the same disease, overlap was limited. Only 36 HPO terms (12.6% of the 285 combined unique terms) were shared between OMIM and Orphanet. OMIM contained 179 terms (62.8%) absent from Orphanet, while Orphanet contained 70 terms (24.6%) absent from OMIM (Figure 1 and Table S1).

**Figure 1.**
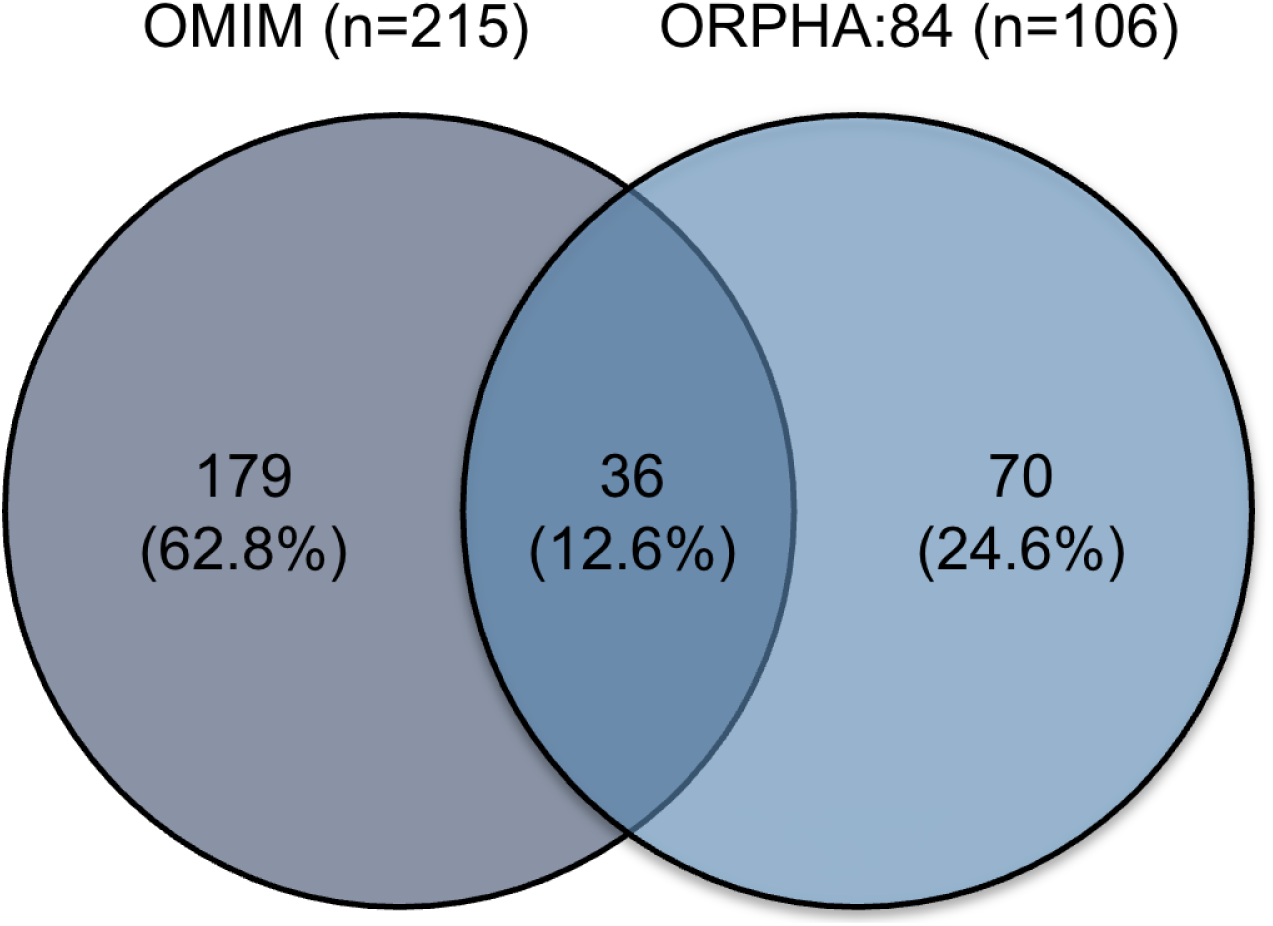
OMIM and Orphanet FA profiles have little overlap. A two-way Venn diagram showing overlap between OMIM (n=215) and Orphanet (n=106) HPO profiles for FA. Only 36 terms are shared (12.6% of 285 combined unique terms).

Anatomical system analysis revealed that both profiles emphasize musculoskeletal (OMIM: 37 terms, 17.2%; Orphanet: 20, 18.9%), nervous system (OMIM: 28, 13.0%; Orphanet: 8, 7.55%), head or neck (OMIM: 19, 8.84%; Orphanet: 15, 14.2%), and genitourinary abnormalities (OMIM: 22, 10.2%; Orphanet: 16, 15.1%). However, several clinically important systems were underrepresented in both profiles, including the immune system (OMIM: 3, 1.40%; Orphanet: 1, 0.94%), integumentary findings (OMIM: 12, 5.58%; Orphanet: 4, 3.77%), digestive system (OMIM: 9, 4.19%; Orphanet: 6, 5.66%), and ear abnormalities (OMIM: 7, 3.26%; Orphanet: 3, 2.83%) (Table 1 and Table S1).

**Table 1.** Distribution of HPO terms by anatomical system across OMIM and Orphanet.

| **Anatomical system** | **OMIM**<br>**(n=215)** | **Orphanet**<br>**(n=106)** |
| --- | --- | --- |
| Musculoskeletal system | 37 (17.2%) | 20 (18.9%) |
| Nervous system | 28 (13.0%) | 8 (7.55%) |
| Genitourinary system | 22 (10.2%) | 16 (15.1%) |
| Head or neck | 19 (8.84%) | 15 (14.2%) |
| Integument | 12 (5.58%) | 4 (3.77%) |
| Cardiovascular system | 12 (5.58%) | 9 (8.49%) |
| Neoplasm | 11 (5.12%) | 2 (1.89%) |
| Endocrine system | 10 (4.65%) | 2 (1.89%) |
| Blood and blood-forming tissues | 10 (4.65%) | 4 (3.77%) |
| Eye | 9 (4.19%) | 10 (9.43%) |
| Digestive system | 9 (4.19%) | 6 (5.66%) |
| Ear | 7 (3.26%) | 3 (2.83%) |
| Metabolism/homeostasis | 7 (3.26%) | 1 (0.94%) |
| Growth abnormality | 7 (3.26%) | 4 (3.77%) |
| Respiratory system | 4 (1.86%) | 0 (0.00%) |
| Connective tissue | 3 (1.40%) | 0 (0.00%) |
| Immune system | 3 (1.40%) | 1 (0.94%) |
| Prenatal development or birth | 3 (1.40%) | 1 (0.94%) |
| Musculature | 2 (0.93%) | 0 (0.00%) |

### A comprehensive custom profile from clinical care guidelines

Our custom FA profile derived from the complete FCF Clinical Care Guidelines comprised 264 unique HPO terms after deduplication and filtering. This profile is the largest HPO-based phenotypic characterization of FA to date, exceeding the combined OMIM profile (215 terms) and the Orphanet profile (106 terms). Three-way comparison revealed 27 terms (6.1% of the 446 total unique terms across all profiles) shared across all three profiles, representing a core set of widely recognized FA phenotypes (Figure 2 and Table S1). The custom profile shared 85 terms (32.2% of custom) with OMIM, comprising 58 terms (22.0%) shared exclusively between these two profiles plus the 27 (10.2%) shared across all three. The custom profile also shared 45 terms (17.0% of custom) with Orphanet, comprising 18 terms (6.8%) shared exclusively plus the 27 (10.2%) shared across all three. Notably, 161 terms (61.0% of the custom profile) were novel and present in neither OMIM nor Orphanet, representing a substantial expansion of computably represented phenotypic knowledge for FA (Figure 2 and Table S1).

**Figure 2.**
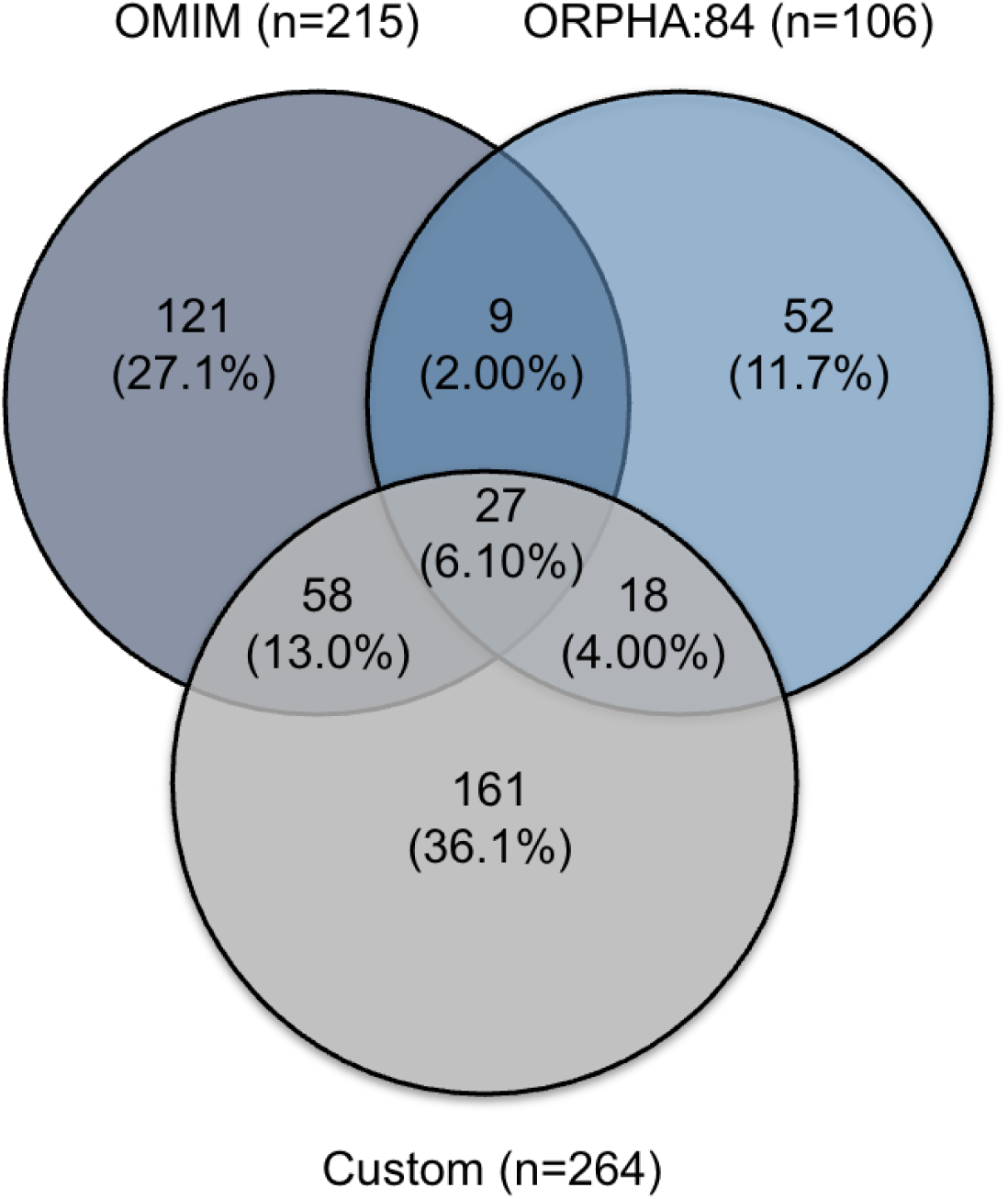
Our custom FA profile expands phenotypic coverage beyond existing OMIM and Orphanet annotations. A three-way Venn diagram comparing OMIM (n=215), Orphanet (n=106), and custom (n=264) HPO profiles for FA. The custom profile contributes 161 novel terms not present in either existing profile.

Anatomical system analysis demonstrated that our custom profile provides expanded coverage across nearly every organ system compared to both existing profiles (Table 2 and Table S1). The most substantial gains were observed in the musculoskeletal system (custom: 66 terms vs. OMIM: 37, Orphanet: 20), genitourinary system (40 vs. 22, 16), digestive system (22 vs. 9, 6), head or neck (20 vs. 19, 15), neoplasms (15 vs. 11, 2), ear abnormalities (12 vs. 7, 3), the immune system (6 vs. 3, 1), and metabolism/homeostasis (10 vs. 7, 1). The only systems where existing profiles showed comparable or greater coverage were the nervous system (OMIM: 28 vs. custom: 15), blood and blood-forming tissues (OMIM: 10 vs. custom: 5), endocrine system (OMIM: 10 vs. custom: 5), and the eye (Orphanet: 10 vs. custom: 7) (Table 2 and Table S1).

**Table 2.** Distribution of HPO terms by anatomical system across OMIM, Orphanet, and custom FA profiles.

| **Anatomical system** | **Custom**<br>**(n=264)** | **OMIM**<br>**(n=215)** | **Orphanet**<br>**(n=106)** |
| --- | --- | --- | --- |
| Musculoskeletal system | 66 (25.0%) | 37 (17.2%) | 20 (18.9%) |
| Genitourinary system | 40 (15.2%) | 22 (10.2%) | 16 (15.1%) |
| Digestive system | 22 (8.33%) | 9 (4.19%) | 6 (5.66%) |
| Head or neck | 20 (7.58%) | 19 (8.84%) | 15 (14.2%) |
| Neoplasm | 15 (5.68%) | 11 (5.12%) | 2 (1.89%) |
| Nervous system | 15 (5.68%) | 28 (13.0%) | 8 (7.55%) |
| Integument | 13 (4.92%) | 12 (5.58%) | 4 (3.77%) |
| Ear | 12 (4.55%) | 7 (3.26%) | 3 (2.83%) |
| Cardiovascular system | 10 (3.79%) | 12 (5.58%) | 9 (8.49%) |
| Metabolism/homeostasis | 10 (3.79%) | 7 (3.26%) | 1 (0.94%) |
| Growth abnormality | 9 (3.41%) | 7 (3.26%) | 4 (3.77%) |
| Eye | 7 (2.65%) | 9 (4.19%) | 10 (9.43%) |
| Immune system | 6 (2.27%) | 3 (1.4%) | 1 (0.94%) |
| Blood and blood-forming tissues | 5 (1.89%) | 10 (4.65%) | 3 (3.77%) |
| Endocrine system | 5 (1.89%) | 10 (4.65%) | 2 (1.89%) |
| Musculature | 3 (1.14%) | 2 (0.93%) | 0 (0%) |
| Connective tissue | 2 (0.76%) | 3 (1.4%) | 0 (0%) |
| Respiratory system | 2 (0.76%) | 4 (1.86%) | 0 (0%) |
| Prenatal development or birth | 1 (0.38%) | 3 (1.4%) | 1 (0.94%) |
| Other/unclassified | 1 (0.38%) | 0 (0%) | 0 (0%) |

### Characterization of novel terms

Analysis of the 161 novel terms unique to the custom profile by anatomical system revealed that the greatest contributions were in areas particularly relevant to FA clinical management (Table 3 and Table S1). Musculoskeletal abnormalities accounted for 45 novel terms (27.9%), including findings such as *Sprengel anomaly* (HP:0000912), *Abnormal rib morphology* (HP:0000772), *Microdontia* (HP:0000691), and *Tooth malposition* (HP:0006482) that are recognized in clinical practice but absent from existing reference profiles. Genitourinary abnormalities were substantially expanded with 25 novel terms (15.5%), capturing phenotypes such as *Testicular atrophy* (HP:0000029), *Chordee* (HP:0000041), *Renal dysplasia* (HP:0000110), *Amenorrhea* (HP:0000141), and *Gonadal dysgenesis* (HP:0000133). The digestive system gained 16 novel terms (9.94%), including *Gastroesophageal reflux* (HP:0002020), *Dysphagia* (HP:0002015), *Constipation* (HP:0002019), *Diarrhea* (HP:0002014), and *Nausea* (HP:0002018). Head or neck phenotypes (10 novel terms, 6.21%) included oral manifestations such as *Gingivitis* (HP:0000230), *Xerostomia* (HP:0000217), *Oral ulcer* (HP:0000155), and *Gingival overgrowth* (HP:0000212) that reflect the oral health complications well-documented in FA clinical care. The integument gained 10 novel terms (6.21%), metabolism/homeostasis 9 terms (5.59%), and the nervous system 9 terms (5.59%) including *Depression* (HP:0000716) and *Anxiety* (HP:0000739). Seven novel neoplasm-related terms (4.35%) were added, including *Basal cell carcinoma* (HP:0002671), *Melanoma* (HP:0002861), and *Oropharyngeal squamous cell carcinoma* (HP:0030448), reflecting the comprehensive cancer predisposition phenotypes described in the clinical guidelines. The ear contributed 6 novel terms (3.73%), the cardiovascular and immune systems each contributed 5 terms (3.11%), and growth contributed 4 terms (2.48%). The remaining 10 novel terms were distributed across endocrine (2, 1.24%), musculature (2, 1.24%), respiratory (2, 1.24%), eye (1, 0.62%), connective tissue (1, 0.62%), prenatal/birth (1, 0.62%), and unclassified (1, 0.62%) categories, capturing clinically relevant findings such as *Delayed puberty* (HP:0000823), *Cardiomyopathy* (HP:0001638), *Pulmonary fibrosis* (HP:0002206), and *Keratoconjunctivitis sicca* (HP:0001097) (Table 3 and Table S1).

**Table 3.**
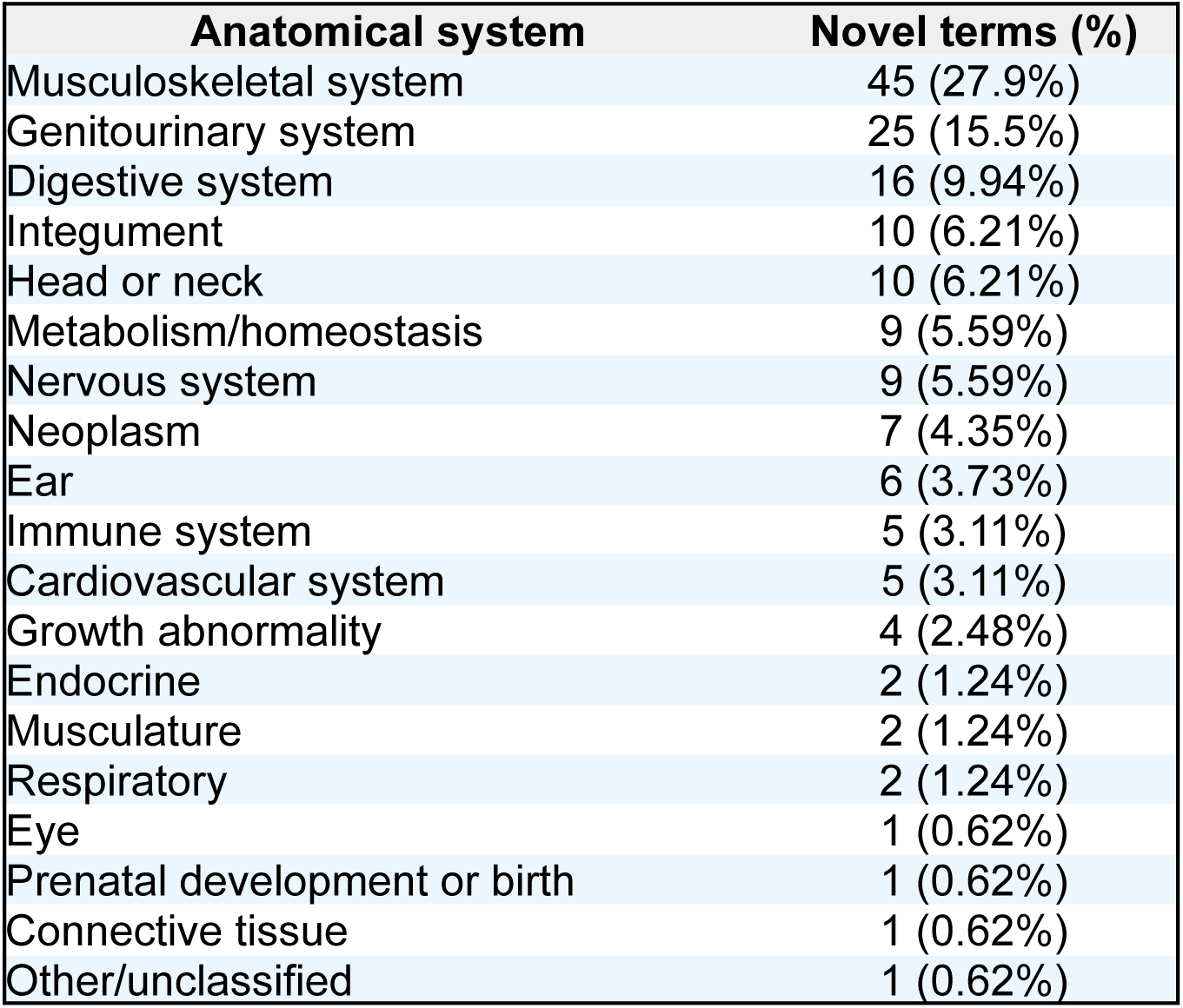
Anatomical system distribution and examples of the 161 novel HPO terms unique to the custom FA profile.

| **Anatomical system** | **Novel terms (%)** |
| --- | --- |
| Musculoskeletal system | 45 (27.9%) |
| Genitourinary system | 25 (15.5%) |
| Digestive system | 16 (9.94%) |
| Integument | 10 (6.21%) |
| Head or neck | 10 (6.21%) |
| Metabolism/homeostasis | 9 (5.59%) |
| Nervous system | 9 (5.59%) |
| Neoplasm | 7 (4.35%) |
| Ear | 6 (3.73%) |
| Immune system | 5 (3.11%) |
| Cardiovascular system | 5 (3.11%) |
| Growth abnormality | 4 (2.48%) |
| Endocrine | 2 (1.24%) |
| Musculature | 2 (1.24%) |
| Respiratory | 2 (1.24%) |
| Eye | 1 (0.62%) |
| Prenatal development or birth | 1 (0.62%) |
| Connective tissue | 1 (0.62%) |
| Other/unclassified | 1 (0.62%) |

## Discussion

We present a comprehensive, curated HPO profile for FA derived from the FCF Clinical Care Guidelines. With 264 HPO terms, this profile represents the most extensive phenotypic characterization of FA in a standardized ontological framework to date. The identification of 161 novel terms not present in either OMIM or Orphanet demonstrates that clinical practice guidelines contain substantial phenotypic knowledge that has not been captured by existing disease annotation resources.

The poor concordance between OMIM and Orphanet FA profiles (only 36 shared terms, 12.6% overlap) underscores a well-recognized challenge in rare disease informatics. This discordance arises from fundamental differences in curation philosophy: OMIM annotates individual genetic subtypes from published case reports, producing 22 separate FA entries of highly variable depth, while Orphanet maintains a single disease-level profile based on expert consensus. Neither approach is inherently superior, but their limited overlap means that downstream applications relying on a single source may miss clinically relevant phenotypes. Our custom profile provides a complementary perspective grounded in the synthesized clinical expertise of FA specialists.

The predominance of novel terms in the musculoskeletal, genitourinary, and limb systems reflects the granularity of phenotypic description in clinical guidelines compared to database annotations. For example, while existing profiles capture broad categories such as skeletal anomalies, the clinical guidelines describe specific findings like *Sprengel anomaly* (HP:0000912), *Osteoporosis* (HP:0000939), and *Arachnodactyly* (HP:0001166) that FA clinicians observe in practice. Similarly, the oral health phenotypes (*Gingivitis* (HP:0000230), *Xerostomia* (HP:0000217), *Microdontia* (HP:0000691)) captured by our profile are well-documented complications of FA but were absent from both OMIM and Orphanet. Our profile also captured mental health phenotypes such as *Anxiety* (HP:0000739) and *Depression* (HP:0000716), reflecting the psychosocial burden emphasized in the clinical guidelines but underrepresented in existing disease annotations [21,22]. These granular phenotypes are precisely the type of clinical observations that could facilitate identification of undiagnosed individuals with FA in EHR data.

This work was made possible by OntoGPT [18], an LLM-based ontological extraction tool developed within the Monarch Initiative ecosystem, that enabled systematic processing of the complete clinical guidelines text. While manual curation of disease-specific HPO profiles is labor-intensive and requires domain expertise, LLM-assisted extraction can accelerate the initial identification of candidate phenotypic terms from clinical knowledge sources. The subsequent manual review step remains essential to ensure clinical relevance and accuracy, but the overall workflow substantially reduces the barrier to developing comprehensive disease-specific profiles. This approach is generalizable to other rare diseases with established clinical practice guidelines.

Several limitations should be noted. First, the custom profile was derived from a single source, the FCF Clinical Care Guidelines, and may not capture phenotypes described only in individual case reports or complementation group-specific studies. Second, while OntoGPT and CurateGPT [18,19] facilitated the extraction and mapping process, the final profile reflects the judgment of the curators in determining clinical relevance of extracted terms. Third, some FA-associated phenotypes lack precise HPO terms, requiring mapping to broader parent terms and representing opportunities for HPO vocabulary expansion through term requests. Finally, the profile has not yet been validated against positive and negative control cohorts of FA, which is a priority for future work.

Future directions include validation of the custom profile against confirmed FA patient cohorts using semantic similarity metrics [15] to quantify its diagnostic utility compared to existing profiles. The profile will also serve as the foundation for EHR-based computable phenotyping of FA, leveraging mappings between clinical terminologies (ICD, SNOMED CT, LOINC) and HPO to identify candidate individuals in large clinical datasets such as the *All of Us* research Program, Truveta, and EPIC Cosmos [23–25]. Integration of this profile into the Monarch Initiative knowledge graph [11] would make it available for phenotype-driven diagnostic applications such as Exomiser and the Monarch API, extending its utility beyond EHR-based phenotyping to genomic variant prioritization and cross-species phenotype analysis. Community feedback on the profile with FA clinicians would further strengthen its clinical validity, with additional specificity of which phenotypes most associate with different complementation groups. Additionally, the approach of extracting HPO profiles from clinical practice guidelines using LLM-assisted tools could be applied to other rare diseases to systematically address gaps in existing phenotype annotations.

Clinical practice guidelines represent a rich and underutilized source of phenotypic knowledge for rare diseases. Our curated HPO profile for Fanconi anemia, comprising 264 terms including 161 not present in OMIM or Orphanet, substantially expands the computably represented phenotypic characterization of this disease. The profile is publicly available and provides a foundation for improved clinical decision support, EHR-based patient identification, and computational phenotyping approaches that can shorten the diagnostic odyssey for individuals with FA, a critical need given that delayed diagnosis directly impacts FA outcomes. More broadly, the LLM-assisted curation approach demonstrated here offers a scalable, reproducible framework that can be applied to any rare disease, helping to close the gap between clinical knowledge and computable disease representations across the rare disease landscape.

## Data Availability

The complete custom HPO profile for FA, including all HPO terms with identifiers and anatomical system classifications, is available as Table S1 and on GitHub [26].

https://github.com/ehurwitz/FA-custom-profile

