## Supplementary material for "A custom phenotypic profile for Fanconi anemia: Addressing gaps in existing disease annotations": Table S1

**Table S1. HPO terms in OMIM, Orphanet, and custom profiles and their corresponding anatomical system.**

| HPO ID | Term label | OMIM | Orphanet | Custom | Anatomical system |
|---|---|---|---|---|---|
| HP:0000010 | *Recurrent urinary tract infections* | | X | | Genitourinary system |
| HP:0000013 | *Hypoplasia of the uterus* | | | X | Genitourinary system |
| HP:0000023 | *Inguinal hernia* | X | | | Digestive system |
| HP:0000027 | *Azoospermia* | | X | X | Genitourinary system |
| HP:0000028 | *Cryptorchidism* | X | X | X | Genitourinary system |
| HP:0000029 | *Testicular atrophy* | | | X | Genitourinary system |
| HP:0000035 | *Abnormal testis morphology* | | X | | Genitourinary system |
| HP:0000041 | *Chordee* | | | X | Genitourinary system |
| HP:0000047 | *Hypospadias* | | X | X | Genitourinary system |
| HP:0000054 | *Micropenis* | X | | | Genitourinary system |
| HP:0000062 | *Ambiguous genitalia* | X | | | Genitourinary system |
| HP:0000072 | *Hydroureter* | | X | X | Genitourinary system |
| HP:0000075 | *Renal duplication* | X | | | Genitourinary system |
| HP:0000076 | *Vesicoureteral reflux* | X | | | Genitourinary system |
| HP:0000077 | *Abnormality of the kidney* | | | X | Genitourinary system |
| HP:0000079 | *Abnormality of the urinary system* | | X | | Genitourinary system |
| HP:0000081 | *Duplicated collecting system* | X | | | Genitourinary system |
| HP:0000083 | *Renal insufficiency* | | X | | Genitourinary system |
| HP:0000085 | *Horseshoe kidney* | X | | X | Genitourinary system |
| HP:0000086 | *Ectopic kidney* | X | | X | Genitourinary system |
| HP:0000089 | *Renal hypoplasia* | X | | X | Genitourinary system |
| HP:0000104 | *Renal agenesis* | X | | X | Genitourinary system |
| HP:0000107 | *Renal cyst* | X | | | Genitourinary system |
| HP:0000110 | *Renal dysplasia* | | | X | Genitourinary system |
| HP:0000122 | *Unilateral renal agenesis* | X | | | Genitourinary system |
| HP:0000125 | *Pelvic kidney* | X | | | Genitourinary system |
| HP:0000126 | *Hydronephrosis* | X | | X | Genitourinary system |
| HP:0000130 | *Abnormality of the uterus* | | X | | Genitourinary system |
| HP:0000132 | *Menorrhagia* | | | X | Genitourinary system |
| HP:0000133 | *Gonadal dysgenesis* | | | X | Genitourinary system |
| HP:0000135 | *Hypogonadism* | X | X | | Endocrine system |
| HP:0000140 | *Abnormality of the menstrual cycle* | | | X | Genitourinary system |
| HP:0000141 | *Amenorrhea* | | | X | Genitourinary system |
| HP:0000144 | *Decreased fertility* | | | X | Genitourinary system |
| HP:0000148 | *Vaginal atresia* | | | X | Genitourinary system |
| HP:0000151 | *Aplasia of the uterus* | X | | X | Genitourinary system |
| HP:0000155 | *Oral ulcer* | | | X | Head or neck |
| HP:0000175 | *Cleft palate* | X | X | X | Head or neck |

| | | | | | |
|---|---|---|---|---|---|
| HP:0000189 | *Narrow palate* | X | | | Head or neck |
| HP:0000212 | *Gingival overgrowth* | | | X | Head or neck |
| HP:0000215 | *Thick upper lip vermilion* | X | | | Head or neck |
| HP:0000217 | *Xerostomia* | | | X | Head or neck |
| HP:0000218 | *High palate* | | X | X | Head or neck |
| HP:0000230 | *Gingivitis* | | | X | Head or neck |
| HP:0000238 | *Hydrocephalus* | X | X | X | Nervous system |
| HP:0000252 | *Microcephaly* | X | X | X | Head or neck |
| HP:0000268 | *Dolichocephaly* | | X | | Musculoskeletal system |
| HP:0000280 | *Coarse facial features* | X | | | Head or neck |
| HP:0000286 | *Epicanthus* | X | X | X | Head or neck |
| HP:0000294 | *Low anterior hairline* | X | | | Head or neck |
| HP:0000307 | *Pointed chin* | | | X | Head or neck |
| HP:0000316 | *Hypertelorism* | X | X | X | Eye |
| HP:0000324 | *Facial asymmetry* | | X | | Head or neck |
| HP:0000325 | *Triangular face* | X | | X | Head or neck |
| HP:0000340 | *Sloping forehead* | | X | | Head or neck |
| HP:0000347 | *Micrognathia* | X | X | X | Musculoskeletal system |
| HP:0000358 | *Posteriorly rotated Ears* | | | X | Ear |
| HP:0000364 | *Hearing abnormality* | | X | | Ear |
| HP:0000365 | *Hearing impairment* | X | X | X | Ear |
| HP:0000369 | *Low-set ears* | X | | X | Ear |
| HP:0000377 | *Abnormal pinna morphology* | | X | X | Ear |
| HP:0000396 | *Overfolded helix* | X | | | Ear |
| HP:0000402 | *Stenosis of the external auditory canal* | | | X | Ear |
| HP:0000405 | *Conductive hearing impairment* | X | | | Ear |
| HP:0000413 | *Atresia of the external auditory canal* | X | | X | Ear |
| HP:0000414 | *Bulbous nose* | X | | | Head or neck |
| HP:0000426 | *Prominent nasal bridge* | X | | | Head or neck |
| HP:0000430 | *Underdeveloped nasal alae* | X | | | Head or neck |
| HP:0000431 | *Wide nasal bridge* | X | | | Head or neck |
| HP:0000437 | *Depressed nasal tip* | X | | | Head or neck |
| HP:0000452 | *Choanal stenosis* | X | | | Head or neck |
| HP:0000453 | *Choanal atresia* | | X | | Head or neck |
| HP:0000463 | *Anteverted nares* | X | | | Head or neck |
| HP:0000465 | *Webbed neck* | X | | | Integument |
| HP:0000470 | *Short neck* | X | | | Musculoskeletal system |
| HP:0000478 | *Abnormality of the eye* | | X | | Eye |
| HP:0000482 | *Microcornea* | X | | X | Eye |
| HP:0000483 | *Astigmatism* | X | X | | Eye |
| HP:0000486 | *Strabismus* | X | X | X | Eye |
| HP:0000492 | *Abnormal eyelid morphology* | | X | | Head or neck |
| HP:0000504 | *Abnormality of vision* | | X | | Eye |
| HP:0000505 | *Visual impairment* | | X | | Eye |
| HP:0000506 | *Telecanthus* | | | X | Head or neck |

| | | | | | |
|---|---|---|---|---|---|
| HP:0000508 | *Ptosis* | | X | X | Head or neck |
| HP:0000518 | *Cataract* | | X | X | Eye |
| HP:0000520 | *Proptosis* | | X | | Head or neck |
| HP:0000527 | *Long eyelashes* | X | | | Integument |
| HP:0000543 | *Optic disc pallor* | X | | | Eye |
| HP:0000545 | *Myopia* | X | | | Eye |
| HP:0000568 | *Microphthalmia* | X | X | X | Eye |
| HP:0000581 | *Blepharophimosis* | X | | | Head or neck |
| HP:0000582 | *Upslanted palpebral fissure* | X | X | | Head or neck |
| HP:0000598 | *Abnormality of the ear* | | | X | Ear |
| HP:0000601 | *Hypotelorism* | X | | X | Eye |
| HP:0000609 | *Optic nerve hypoplasia* | X | | | Nervous system |
| HP:0000639 | *Nystagmus* | | X | | Eye |
| HP:0000689 | *Dental malocclusion* | X | | | Musculoskeletal system |
| HP:0000691 | *Microdontia* | | | X | Musculoskeletal system |
| HP:0000692 | *Tooth malposition* | | | X | Musculoskeletal system |
| HP:0000696 | *Delayed eruption of permanent teeth* | | | X | Musculoskeletal system |
| HP:0000704 | *Periodontitis* | | | X | Head or neck |
| HP:0000707 | *Abnormality of the nervous system* | | | X | Nervous system |
| HP:0000716 | *Depression* | | | X | Nervous system |
| HP:0000739 | *Anxiety* | | | X | Nervous system |
| HP:0000750 | *Delayed speech and language development* | X | | | Nervous system |
| HP:0000772 | *Abnormal rib morphology* | | | X | Musculoskeletal system |
| HP:0000789 | *Infertility* | | | X | Genitourinary system |
| HP:0000798 | *Oligozoospermia* | | | X | Genitourinary system |
| HP:0000813 | *Bicornuate uterus* | | X | X | Genitourinary system |
| HP:0000815 | *Hypergonadotropic hypogonadism* | X | | | Endocrine system |
| HP:0000819 | *Diabetes mellitus* | | | X | Metabolism/homeostasis |
| HP:0000821 | *Hypothyroidism* | X | | X | Endocrine system |
| HP:0000823 | *Delayed puberty* | | | X | Endocrine system |
| HP:0000824 | *Decreased response to growth hormone stimulation test* | X | | | Endocrine system |
| HP:0000855 | *Insulin resistance* | | | X | Metabolism/homeostasis |
| HP:0000858 | *Irregular menstruation* | | | X | Genitourinary system |
| HP:0000864 | *Abnormality of the hypothalamus-pituitary axis* | | X | | Endocrine system |
| HP:0000868 | *Decreased fertility in females* | | | X | Genitourinary system |
| HP:0000876 | *Oligomenorrhea* | | | X | Genitourinary system |
| HP:0000902 | *Rib fusion* | X | | | Musculoskeletal system |
| HP:0000912 | *Sprengel anomaly* | | | X | Musculoskeletal system |
| HP:0000924 | *Abnormality of the skeletal system* | | | X | Musculoskeletal system |
| HP:0000939 | *Osteoporosis* | | | X | Musculoskeletal system |

| | | | | | |
|---|---|---|---|---|---|
| HP:0000953 | *Hyperpigmentation of the skin* | X | | X | Integument |
| HP:0000957 | *Cafe-au-lait spot* | X | | X | Integument |
| HP:0000958 | *Dry skin* | | | X | Integument |
| HP:0000960 | *Sacral dimple* | X | | | Integument |
| HP:0000967 | *Petechiae* | | | X | Integument |
| HP:0000978 | *Bruising susceptibility* | X | | X | Blood and blood-forming tissues |
| HP:0000979 | *Purpura* | | | X | Integument |
| HP:0001000 | *Abnormality of skin pigmentation* | X | X | X | Integument |
| HP:0001010 | *Hypopigmentation of the skin* | | | X | Integument |
| HP:0001017 | *Anemic pallor* | X | | | Integument |
| HP:0001045 | *Vitiligo* | X | | | Integument |
| HP:0001053 | *Hypopigmented skin patches* | | X | | Integument |
| HP:0001072 | *Thickened skin* | | | X | Integument |
| HP:0001097 | *Keratoconjunctivitis sicca* | | | X | Eye |
| HP:0001155 | *Abnormality of the hand* | | | X | Musculoskeletal system |
| HP:0001156 | *Brachydactyly* | | | X | Musculoskeletal system |
| HP:0001159 | *Syndactyly* | X | | | Musculoskeletal system |
| HP:0001166 | *Arachnodactyly* | | | X | Musculoskeletal system |
| HP:0001172 | *Abnormal thumb morphology* | X | X | X | Musculoskeletal system |
| HP:0001177 | *Preaxial hand polydactyly* | X | | X | Musculoskeletal system |
| HP:0001195 | *Single umbilical artery* | X | | | Prenatal development or birth |
| HP:0001199 | *Triphalangeal thumb* | | X | X | Musculoskeletal system |
| HP:0001233 | *2-3 finger cutaneous syndactyly* | X | | | Musculoskeletal system |
| HP:0001238 | *Slender finger* | | | X | Musculoskeletal system |
| HP:0001245 | *Small thenar eminence* | X | | X | musculature |
| HP:0001249 | *Intellectual disability* | X | X | | Nervous system |
| HP:0001251 | *Ataxia* | X | | | Nervous system |
| HP:0001252 | *Hypotonia* | X | | | musculature |
| HP:0001263 | *Global developmental delay* | X | X | | Nervous system |
| HP:0001273 | *Abnormal corpus callosum morphology* | | | X | Nervous system |
| HP:0001274 | *Agenesis of corpus callosum* | X | | | Nervous system |
| HP:0001321 | *Cerebellar hypoplasia* | X | | | Nervous system |
| HP:0001328 | *Specific learning disability* | X | | | Nervous system |
| HP:0001331 | *Absent septum pellucidum* | X | | X | Nervous system |
| HP:0001347 | *Hyperreflexia* | | X | X | Nervous system |
| HP:0001360 | *Holoprosencephaly* | | | X | Nervous system |
| HP:0001363 | *Craniosynostosis* | | | X | Musculoskeletal system |
| HP:0001371 | *Flexion contracture* | X | | | Connective tissue |
| HP:0001374 | *Congenital hip dislocation* | | | X | Musculoskeletal system |
| HP:0001385 | *Hip dysplasia* | | | X | Musculoskeletal system |
| HP:0001392 | *Abnormality of the liver* | | X | | Digestive system |
| HP:0001498 | *Carpal bone hypoplasia* | | | X | Musculoskeletal system |
| HP:0001508 | *Failure to thrive* | X | | X | Growth abnormality |

| | | | | | |
|---|---|---|---|---|---|
| HP:0001510 | *Growth delay* | X | X | X | Growth abnormality |
| HP:0001511 | *Intrauterine growth retardation* | X | X | X | Growth abnormality |
| HP:0001513 | *Obesity* | | | X | Growth abnormality |
| HP:0001518 | *Small for gestational age* | X | | X | Growth abnormality |
| HP:0001537 | *Umbilical hernia* | | X | | Digestive system |
| HP:0001545 | *Anteriorly placed anus* | X | | | Digestive system |
| HP:0001561 | *Polyhydramnios* | X | | | Prenatal development or birth |
| HP:0001562 | *Oligohydramnios* | | X | | Prenatal development or birth |
| HP:0001572 | *Macrodontia* | X | | | Musculoskeletal system |
| HP:0001627 | *Abnormal heart morphology* | X | | | Cardiovascular system |
| HP:0001629 | *Ventricular septal defect* | X | | X | Cardiovascular system |
| HP:0001631 | *Atrial septal defect* | X | X | X | Cardiovascular system |
| HP:0001636 | *Tetralogy of Fallot* | X | X | X | Cardiovascular system |
| HP:0001638 | *Cardiomyopathy* | | | X | musculature |
| HP:0001639 | *Hypertrophic cardiomyopathy* | | X | | Cardiovascular system |
| HP:0001642 | *Pulmonic stenosis* | | | X | Cardiovascular system |
| HP:0001643 | *Patent ductus arteriosus* | X | X | X | Cardiovascular system |
| HP:0001646 | *Abnormal aortic valve morphology* | | X | | Cardiovascular system |
| HP:0001650 | *Aortic valve stenosis* | | | X | Cardiovascular system |
| HP:0001651 | *Dextrocardia* | X | | | Cardiovascular system |
| HP:0001655 | *Patent foramen ovale* | X | | | Cardiovascular system |
| HP:0001662 | *Bradycardia* | X | | | Cardiovascular system |
| HP:0001671 | *Abnormal cardiac septum morphology* | | X | | Cardiovascular system |
| HP:0001674 | *Complete atrioventricular canal defect* | X | | | Cardiovascular system |
| HP:0001679 | *Abnormal aortic morphology* | | X | | Cardiovascular system |
| HP:0001680 | *Coarctation of aorta* | X | | X | Cardiovascular system |
| HP:0001734 | *Annular pancreas* | X | | X | Endocrine system |
| HP:0001741 | *Phimosis* | | | X | Genitourinary system |
| HP:0001748 | *Polysplenia* | X | | | Immune system |
| HP:0001760 | *Abnormal foot morphology* | | X | | Musculoskeletal system |
| HP:0001762 | *Talipes equinovarus* | X | | X | Connective tissue |
| HP:0001763 | *Pes planus* | | X | | Musculoskeletal system |
| HP:0001770 | *Toe syndactyly* | | X | | Musculoskeletal system |
| HP:0001776 | *Bilateral talipes equinovarus* | X | | | Connective tissue |
| HP:0001824 | *Weight loss* | | X | | Growth abnormality |
| HP:0001864 | *Clinodactyly of the 5th toe* | X | | | Musculoskeletal system |
| HP:0001871 | *Abnormality of blood and blood-forming tissues* | | X | | Blood and blood-forming tissues |
| HP:0001873 | *Thrombocytopenia* | X | X | X | Blood and blood-forming tissues |
| HP:0001875 | *Decreased total neutrophil count* | X | | X | Immune system |
| HP:0001876 | *Pancytopenia* | X | | | Blood and blood-forming tissues |
| HP:0001882 | *Decreased total leukocyte count* | X | X | | Immune system |

| | | | | | |
|---|---|---|---|---|---|
| HP:0001896 | *Reticulocytopenia* | X | | | Blood and blood-forming tissues |
| HP:0001903 | *Anemia* | X | X | X | Blood and blood-forming tissues |
| HP:0001909 | *Leukemia* | X | | X | Neoplasm |
| HP:0001915 | *Aplastic anemia* | X | | X | Blood and blood-forming tissues |
| HP:0001963 | *Abnormal speech discrimination* | | | X | Ear |
| HP:0002007 | *Frontal bossing* | | X | X | Head or neck |
| HP:0002014 | *Diarrhea* | | | X | Digestive system |
| HP:0002015 | *Dysphagia* | | | X | Digestive system |
| HP:0002018 | *Nausea* | | | X | Digestive system |
| HP:0002019 | *Constipation* | | | X | Digestive system |
| HP:0002020 | *Gastroesophageal reflux* | | | X | Digestive system |
| HP:0002023 | *Anal atresia* | X | X | X | Digestive system |
| HP:0002024 | *Malabsorption* | | | X | Digestive system |
| HP:0002027 | *Abdominal pain* | | | X | Digestive system |
| HP:0002032 | *Esophageal atresia* | X | | X | Digestive system |
| HP:0002079 | *Hypoplasia of the corpus callosum* | X | | | Nervous system |
| HP:0002089 | *Pulmonary hypoplasia* | X | | | Respiratory system |
| HP:0002090 | *Pneumonia* | X | | | Respiratory system |
| HP:0002101 | *Abnormal lung lobation* | X | | | Respiratory system |
| HP:0002119 | *Ventriculomegaly* | X | X | X | Nervous system |
| HP:0002126 | *Polymicrogyria* | X | | | Nervous system |
| HP:0002144 | *Tethered cord* | X | | | Nervous system |
| HP:0002188 | *Delayed CNS myelination* | X | | | Nervous system |
| HP:0002206 | *Pulmonary fibrosis* | | | X | Respiratory system |
| HP:0002245 | *Meckel diverticulum* | | X | | Digestive system |
| HP:0002247 | *Duodenal atresia* | X | | X | Digestive system |
| HP:0002251 | *Aganglionic megacolon* | | X | | Nervous system |
| HP:0002308 | *Chiari malformation* | X | | X | Nervous system |
| HP:0002414 | *Spina bifida* | | X | X | Musculoskeletal system |
| HP:0002518 | *Abnormal periventricular white matter morphology* | X | | | Nervous system |
| HP:0002566 | *Intestinal malrotation* | | | X | Digestive system |
| HP:0002575 | *Tracheoesophageal fistula* | X | X | X | Digestive system |
| HP:0002578 | *Gastroparesis* | | | X | musculature |
| HP:0002607 | *Bowel incontinence* | | | X | Digestive system |
| HP:0002650 | *Scoliosis* | X | X | X | Musculoskeletal system |
| HP:0002664 | *Neoplasm* | | X | X | Neoplasm |
| HP:0002667 | *Nephroblastoma* | X | | X | Neoplasm |
| HP:0002671 | *Basal cell carcinoma* | | | X | Neoplasm |
| HP:0002691 | *Platybasia* | | | X | Musculoskeletal system |
| HP:0002718 | *Recurrent bacterial infections* | | | X | Immune system |
| HP:0002719 | *Recurrent infections* | | | X | Immune system |
| HP:0002808 | *Kyphosis* | | | X | Musculoskeletal system |

| | | | | | |
|---|---|---|---|---|---|
| HP:0002814 | *Abnormality of the lower limb* | | | X | Musculoskeletal system |
| HP:0002817 | *Abnormality of the upper limb* | | X | X | Musculoskeletal system |
| HP:0002823 | *Abnormal femur morphology* | | X | | Musculoskeletal system |
| HP:0002827 | *Hip dislocation* | | X | | Musculoskeletal system |
| HP:0002841 | *Recurrent fungal infections* | | | X | Immune system |
| HP:0002860 | *Squamous cell carcinoma* | X | | X | Neoplasm |
| HP:0002861 | *Melanoma* | | | X | Neoplasm |
| HP:0002863 | *Myelodysplasia* | X | X | X | Neoplasm |
| HP:0002885 | *Medulloblastoma* | X | | X | Neoplasm |
| HP:0002949 | *Fused cervical vertebrae* | X | | | Musculoskeletal system |
| HP:0002984 | *Hypoplasia of the radius* | X | | X | Musculoskeletal system |
| HP:0002996 | *Limited elbow movement* | | | X | Musculoskeletal system |
| HP:0003002 | *Breast carcinoma* | X | | | Neoplasm |
| HP:0003006 | *Neuroblastoma* | X | | X | Neoplasm |
| HP:0003022 | *Hypoplasia of the ulna* | | X | | Musculoskeletal system |
| HP:0003031 | *Ulnar bowing* | | | X | Musculoskeletal system |
| HP:0003074 | *Hyperglycemia* | | | X | Metabolism/homeostasis |
| HP:0003119 | *Abnormal circulating lipid concentration* | | | X | Metabolism/homeostasis |
| HP:0003213 | *Deficient excision of UV-induced pyrimidine dimers in DNA* | X | | | Metabolism/homeostasis |
| HP:0003214 | *Prolonged G2 phase of cell cycle* | X | | | Metabolism/homeostasis |
| HP:0003220 | *Abnormality of chromosome stability* | X | X | | Metabolism/homeostasis |
| HP:0003221 | *Chromosomal breakage induced by crosslinking agents* | X | | X | Metabolism/homeostasis |
| HP:0003241 | *External genital hypoplasia* | X | | X | Genitourinary system |
| HP:0003250 | *Aplasia of the vagina* | | | X | Genitourinary system |
| HP:0003251 | *Male infertility* | X | | | Genitourinary system |
| HP:0003254 | *Abnormality of DNA repair* | | | X | Metabolism/homeostasis |
| HP:0003272 | *Abnormal hip bone morphology* | | | X | Musculoskeletal system |
| HP:0003452 | *Increased circulating iron concentration* | | | X | Metabolism/homeostasis |
| HP:0003468 | *Abnormal vertebral morphology* | X | | X | Musculoskeletal system |
| HP:0003764 | *Nevus* | X | | | Integument |
| HP:0003774 | *Stage 5 chronic kidney disease* | X | | | Genitourinary system |
| HP:0003834 | *Shoulder dislocation* | X | | | Musculoskeletal system |
| HP:0003956 | *Bowed forearm bones* | | | X | Musculoskeletal system |
| HP:0003974 | *Absent radius* | X | | X | Musculoskeletal system |
| HP:0003982 | *Aplasia of the ulna* | | | X | Musculoskeletal system |
| HP:0004209 | *Clinodactyly of the 5th finger* | X | X | | Musculoskeletal system |
| HP:0004247 | *Small scaphoid* | | | X | Musculoskeletal system |
| HP:0004253 | *Absent trapezium* | | | X | Musculoskeletal system |
| HP:0004255 | *Small trapezium* | | | X | Musculoskeletal system |
| HP:0004322 | *Short stature* | X | X | X | Growth abnormality |

| | | | | | |
|---|---|---|---|---|---|
| HP:0004323 | *Abnormality of body weight* | | | X | Growth abnormality |
| HP:0004325 | *Decreased body weight* | X | | | Growth abnormality |
| HP:0004349 | *Reduced bone mineral density* | | X | X | Musculoskeletal system |
| HP:0004395 | *Malnutrition* | | | X | Digestive system |
| HP:0004429 | *Recurrent viral infections* | | | X | Immune system |
| HP:0004510 | *Pancreatic islet-cell hyperplasia* | X | | | Endocrine system |
| HP:0004590 | *Hypoplastic sacrum* | X | | X | Musculoskeletal system |
| HP:0004602 | *Cervical C2/C3 vertebral fusion* | | | X | Musculoskeletal system |
| HP:0004712 | *Renal malrotation* | | | X | Genitourinary system |
| HP:0004808 | *Acute myeloid leukemia* | X | | X | Neoplasm |
| HP:0004935 | *Pulmonary artery atresia* | | | X | Cardiovascular system |
| HP:0004977 | *Bilateral radial aplasia* | X | | | Musculoskeletal system |
| HP:0005214 | *Intestinal obstruction* | | | X | Digestive system |
| HP:0005343 | *Hypoplasia of the bladder* | X | | | Genitourinary system |
| HP:0005344 | *Abnormal carotid artery morphology* | | X | | Cardiovascular system |
| HP:0005473 | *Fusion of middle ear ossicles* | | | X | Musculoskeletal system |
| HP:0005518 | *Increased mean corpuscular volume* | X | | | Blood and blood-forming tissues |
| HP:0005522 | *Pyridoxine-responsive sideroblastic anemia* | | X | | Blood and blood-forming tissues |
| HP:0005528 | *Bone marrow hypocellularity* | X | | X | Blood and blood-forming tissues |
| HP:0005632 | *Absent forearm* | X | | | Musculoskeletal system |
| HP:0005709 | *2-3 toe cutaneous syndactyly* | X | | | Musculoskeletal system |
| HP:0005743 | *Avascular necrosis of the capital femoral epiphysis* | | | X | Musculoskeletal system |
| HP:0005792 | *Short humerus* | X | | X | Musculoskeletal system |
| HP:0005912 | *Biliary atresia* | X | | X | Digestive system |
| HP:0006101 | *Finger syndactyly* | | X | | Musculoskeletal system |
| HP:0006190 | *Radially deviated wrists* | | | X | Musculoskeletal system |
| HP:0006248 | *Limited wrist movement* | | | X | Musculoskeletal system |
| HP:0006254 | *Elevated circulating alpha-fetoprotein concentration* | X | | | Metabolism/homeostasis |
| HP:0006265 | *Aplasia/Hypoplasia of fingers* | | X | | Musculoskeletal system |
| HP:0006349 | *Agenesis of permanent teeth* | X | | | Musculoskeletal system |
| HP:0006433 | *Radial ray deficiency* | X | | X | Musculoskeletal system |
| HP:0006482 | *Abnormal dental morphology* | | | X | Musculoskeletal system |
| HP:0006501 | *Aplasia/Hypoplasia of the radius* | | X | X | Musculoskeletal system |
| HP:0006660 | *Aplastic clavicle* | | | X | Musculoskeletal system |
| HP:0006727 | *T-cell acute lymphoblastic leukemias* | X | | | Neoplasm |
| HP:0006824 | *Cranial nerve paralysis* | | X | | Nervous system |
| HP:0007018 | *Attention deficit hyperactivity disorder* | X | | | Nervous system |
| HP:0007099 | *Chiari type I malformation* | X | | | Nervous system |
| HP:0007400 | *Irregular hyperpigmentation* | | X | | Integument |

| | | | | | |
|---|---|---|---|---|---|
| HP:0007565 | *Multiple cafe-au-lait spots* | X | X | | Integument |
| HP:0007587 | *Numerous pigmented freckles* | | | X | Integument |
| HP:0007766 | *Optic disc hypoplasia* | X | | | Eye |
| HP:0007874 | *Almond-shaped palpebral fissure* | | X | X | Head or neck |
| HP:0008053 | *Aplasia/Hypoplasia of the iris* | | X | | Eye |
| HP:0008070 | *Sparse hair* | X | | | Integument |
| HP:0008209 | *Premature ovarian insufficiency* | | | X | Genitourinary system |
| HP:0008551 | *Microtia* | X | | X | Ear |
| HP:0008661 | *Urethral stenosis* | | | X | Genitourinary system |
| HP:0008678 | *Renal hypoplasia/aplasia* | | X | | Genitourinary system |
| HP:0008734 | *Decreased testicular size* | | | X | Genitourinary system |
| HP:0008839 | *Hypoplastic pelvis* | | | X | Musculoskeletal system |
| HP:0008897 | *Postnatal growth retardation* | X | | | Growth abnormality |
| HP:0009592 | *Astrocytoma* | | | X | Neoplasm |
| HP:0009603 | *Deviation of the thumb* | | | X | Musculoskeletal system |
| HP:0009623 | *Proximal placement of thumb* | X | | | Musculoskeletal system |
| HP:0009660 | *Short phalanx of the thumb* | | | X | Musculoskeletal system |
| HP:0009777 | *Absent thumb* | X | | X | Musculoskeletal system |
| HP:0009778 | *Short thumb* | X | | X | Musculoskeletal system |
| HP:0009804 | *Tooth agenesis* | | | X | Musculoskeletal system |
| HP:0009821 | *Forearm undergrowth* | | | X | Musculoskeletal system |
| HP:0009829 | *Phocomelia* | X | | | Musculoskeletal system |
| HP:0009892 | *Anotia* | X | | X | Ear |
| HP:0009942 | *Duplication of thumb phalanx* | X | | X | Musculoskeletal system |
| HP:0009943 | *Complete duplication of thumb phalanx* | X | | | Musculoskeletal system |
| HP:0009944 | *Partial duplication of thumb phalanx* | X | | X | Musculoskeletal system |
| HP:0010034 | *Short 1st metacarpal* | X | | | Musculoskeletal system |
| HP:0010035 | *Aplasia of the 1st metacarpal* | X | | | Musculoskeletal system |
| HP:0010293 | *Aplasia/Hypoplasia of the uvula* | | X | | Head or neck |
| HP:0010305 | *Absence of the sacrum* | | | X | Musculoskeletal system |
| HP:0010442 | *Polydactyly* | | | X | Musculoskeletal system |
| HP:0010445 | *Primum atrial septal defect* | X | | | Cardiovascular system |
| HP:0010461 | *Abnormality of the male genitalia* | | | X | Genitourinary system |
| HP:0010469 | *Absent testis* | | X | X | Genitourinary system |
| HP:0010628 | *Facial palsy* | X | | X | Nervous system |
| HP:0010664 | *Fusion of the left and right thalami* | X | | | Nervous system |
| HP:0010704 | *1-2 finger cutaneous syndactyly* | | | X | Musculoskeletal system |
| HP:0011014 | *Abnormal glucose homeostasis* | | | X | Metabolism/homeostasis |
| HP:0011069 | *Supernumerary tooth* | | | X | Musculoskeletal system |
| HP:0011107 | *Recurrent aphthous stomatitis* | | | X | Head or neck |
| HP:0011109 | *Chronic sinusitis* | | | X | Respiratory system |

| | | | | | |
|---|---|---|---|---|---|
| HP:0011133 | *Increased sensitivity to ionizing radiation* | | | X | Metabolism/homeostasis |
| HP:0011419 | *Placental abruption* | X | | | Prenatal development or birth |
| HP:0011590 | *Double aortic arch* | | | X | Cardiovascular system |
| HP:0011800 | *Midface retrusion* | X | | X | Head or neck |
| HP:0011834 | *Moyamoya phenomenon* | | | X | Nervous system |
| HP:0011835 | *Absent scaphoid* | X | | X | Musculoskeletal system |
| HP:0011940 | *Anterior wedging of T12* | X | | | Musculoskeletal system |
| HP:0011968 | *Feeding difficulties* | X | | X | Digestive system |
| HP:0012041 | *Decreased fertility in males* | | X | | Genitourinary system |
| HP:0012165 | *Oligodactyly* | X | | | Musculoskeletal system |
| HP:0012174 | *Glioblastoma multiforme* | | | X | Neoplasm |
| HP:0012182 | *Oropharyngeal squamous cell carcinoma* | | | X | Neoplasm |
| HP:0012210 | *Abnormal renal morphology* | X | X | X | Genitourinary system |
| HP:0012285 | *Abnormal hypothalamus physiology* | | | X | Endocrine system |
| HP:0012506 | *Small pituitary gland* | X | | | Endocrine system |
| HP:0012622 | *Chronic kidney disease* | | | X | Genitourinary system |
| HP:0012639 | *Abnormal nervous system morphology* | | X | | Nervous system |
| HP:0012745 | *Short palpebral fissure* | X | X | X | Head or neck |
| HP:0012799 | *Unilateral facial palsy* | X | | | Nervous system |
| HP:0020073 | *Hypopigmented macule* | X | | | Integument |
| HP:0020128 | *Aplasia of the olfactory tract* | X | | | Nervous system |
| HP:0025023 | *Rectal atresia* | X | | | Digestive system |
| HP:0025031 | *Abnormality of the digestive system* | | | X | Digestive system |
| HP:0025127 | *Actinic keratosis* | | | X | Integument |
| HP:0025261 | *Stiff finger* | | | X | Musculoskeletal system |
| HP:0025318 | *Ovarian carcinoma* | X | | | Neoplasm |
| HP:0025474 | *Erythematous plaque* | | | X | Integument |
| HP:0025502 | *Overweight* | | | X | Growth abnormality |
| HP:0030048 | *Colpocephaly* | X | | | Nervous system |
| HP:0030079 | *Cervix cancer* | | | X | Neoplasm |
| HP:0030084 | *Clinodactyly* | X | | X | Musculoskeletal system |
| HP:0030260 | *Microphallus* | X | | X | Genitourinary system |
| HP:0030283 | *Partial absence of the septum pellucidum* | X | | | Nervous system |
| HP:0030680 | *Abnormal cardiovascular system morphology* | X | | | Cardiovascular system |
| HP:0030996 | *Megaduodenum* | | | X | Digestive system |
| HP:0031095 | *Abnormal humerus morphology* | | | X | Musculoskeletal system |
| HP:0031640 | *Abnormal radial artery morphology* | | | X | Cardiovascular system |
| HP:0031689 | *Megakaryocyte dysplasia* | X | | | Blood and blood-forming tissues |
| HP:0031703 | *Abnormal ear morphology* | | | X | Ear |
| HP:0031936 | *Delayed ability to walk* | X | | | Nervous system |

| | | | | | |
|---|---|---|---|---|---|
| HP:0031965 | *Increased RBC distribution width* | X | | | Blood and blood-forming tissues |
| HP:0032043 | *Odynophagia* | | | X | Digestive system |
| HP:0032154 | *Aphthous ulcer* | | | X | Head or neck |
| HP:0032188 | *Cellular hypersensitivity to mitomycin C* | X | | | Metabolism/homeostasis |
| HP:0032464 | *Ureteral hypoplasia* | X | | | Genitourinary system |
| HP:0033183 | *Bilobed right lung* | X | | | Respiratory system |
| HP:0033667 | *Diminished mental health* | | | X | Other/unclassified |
| HP:0033725 | *Thin corpus callosum* | X | | | Nervous system |
| HP:0034057 | *Fetal anomaly* | | | X | Prenatal development or birth |
| HP:0034231 | *Sigmoid kidney* | | | X | Genitourinary system |
| HP:0034323 | *Reduced circulating growth hormone concentration* | X | | X | Endocrine system |
| HP:0034585 | *Cochlear nerve hypoplasia* | | | X | Nervous system |
| HP:0034681 | *Finger joint contracture* | | | X | Connective tissue |
| HP:0034976 | *Absent pituitary stalk* | X | | | Endocrine system |
| HP:0040012 | *Chromosome breakage* | X | | | Metabolism/homeostasis |
| HP:0040071 | *Abnormal morphology of ulna* | | X | | Musculoskeletal system |
| HP:0040075 | *Hypopituitarism* | X | | | Endocrine system |
| HP:0040090 | *Abnormal tympanic membrane morphology* | | | X | Ear |
| HP:0040183 | *Encopresis* | | | X | Digestive system |
| HP:0040189 | *Scaling skin* | | | X | Integument |
| HP:0040270 | *Impaired glucose tolerance* | | | X | Metabolism/homeostasis |
| HP:0045005 | *Neural tube defect* | | | X | Nervous system |
| HP:0045025 | *Narrow palpebral fissure* | | | X | Head or neck |
| HP:0100026 | *Arteriovenous malformation* | | X | | Cardiovascular system |
| HP:0100542 | *Abnormal localization of kidney* | | X | | Genitourinary system |
| HP:0100559 | *Lower limb asymmetry* | | | X | Growth abnormality |
| HP:0100587 | *Abnormal preputium morphology* | | X | | Genitourinary system |
| HP:0100615 | *Ovarian Neoplasm* | X | | | Neoplasm |
| HP:0100760 | *Clubbing of toes* | | X | | Musculoskeletal system |
| HP:0100842 | *Septo-optic dysplasia* | X | | | Nervous system |
| HP:0100867 | *Duodenal stenosis* | | X | | Digestive system |
| HP:0200036 | *Skin nodule* | | | X | Integument |
| HP:0200043 | *Verrucae* | | | X | Neoplasm |
| HP:0410028 | *Recurrent oral herpes* | | | X | Immune system |
| HP:5200320 | *Diminishment of relationship seeking* | | | X | Nervous system |
| HP:6000064 | *Excessive eructation* | | | X | Digestive system |
| HP:6000942 | *Thumb hypoplasia grade 4* | | | X | Musculoskeletal system |
